# Personalization and network specificity of cerebellar TMS in schizophrenia

**DOI:** 10.64898/2025.12.19.25342404

**Authors:** Emma Joncas, Emily Payne, Eunice Lee, Isabella Demo, Madelaine Nye, Virginie-Anne Chouinard, Moritz Dannhauer, Roscoe O. Brady, Mark A. Halko, Rujuta U Parlikar

**Affiliations:** Schizophrenia and Bipolar Research Program, McLean Hospital, Belmont MA, and Harvard Medical School, Boston, MA; Psychiatry, Beth Israel Deaconess Medical Center, Boston, MA, and Harvard Medical School, Boston, MA; Alliance for Brain Stimulation, Department for Computer Science, East Carolina University, Greenville, NC

## Abstract

**Background:** Cerebellar transcranial magnetic stimulation (TMS) may serve as an adjuvant therapy for psychosis symptoms, most recently we have shown improvements in negative symptoms. Historically, cerebellum TMS has not utilized functional neuroanatomy for targeting, and the precision of TMS to the cerebellum is unclear. A classical view of the cerebellum as solely involved in motor computations has been updated with the discovery of rich non-motor connectivity including the default, dorsal attention, frontoparietal control and ventral attention networks. We sought to assess cerebellar TMS magnetic field effect within individually defined networks of the cerebellum.

**Methods:** Imaging data from schizophrenia and schizoaffective participants (n=27) in a double-blinded trial of cerebellar TMS (NCT05343598) was used. Individualized resting-state connectivity fMRI maps of the cerebellum was computed for 7 canonical networks (Yeo et al 2011; Buckner et al 2011). Individualized TMS simulations were computed in SimNIBS with real-world participant-specific coil placement and intensity determination.

**Results:** The peak stimulation effect (99th percentile) for each network in each participant was computed. The electric field induced by cerebellar TMS predominantly engaged specific functional networks more than others (p<0.001), indicating selective targeting of these networks. The strongest effects were found on default (44.4%), limbic (37%) and frontoparietal control (11.1%) networks. Cerebellar brain network organization was found to be similar in the patient sample to previously published large-sample organization.

**Conclusions:** For personalized TMS, it is important to consider the targeted network, as well as the potential off-target network effects. Our findings demonstrate that cerebellar TMS has the strongest field effect on non-motor, cognitive and affective networks within the cerebellum. These results suggest cerebellar TMS may be ideal for schizophrenia symptoms unaddressed by pharmacological treatments, and effects may vary by individual network topology.

## INTRODUCTION

The classical model of the cerebellum having a critical role only in motor function has seen a large-scale revision based on neuroimaging findings. Brain networks associated with the cerebral cortex are found in the cerebellum [1], with some estimates suggesting that only 20-30% of the cerebellum connects to the motor and somatosensory cortex [2].

Interest in the cerebellum’s role in schizophrenia was renewed with seminal work from Andreasen [3, 4], and Schmahmann [5, 6]. Imaging studies have linked cerebellar function and dysfunction to symptoms of schizophrenia [7]. This has been further reinforced by cerebellum-targeted therapies [8, 9] which have shown clinical improvement in patients with psychosis [10–12].

Transcranial magnetic stimulation (TMS) is a non-invasive brain stimulation technique that is based on the principle that dynamically modulating magnetic fields results in the generation of an electric field in the underlying cortex through a handheld coil. TMS has been widely used both in investigational [13] and therapeutic capacities [14, 15] for understanding and treating neuropsychiatric conditions. Given the growing body of evidence supporting the pathophysiological role of the cerebellum in schizophrenia, cerebellar TMS has been gaining substantial torque as a therapeutic intervention.

A growing body of literature has found merits in targeting cerebellar regions and networks using TMS [16]. However, there is substantial heterogeneity in the clinical outcomes in patients with schizophrenia compared to evidence backing TMS application in depression [17]. Several controlled trials were conducted to further elucidate these findings [11, 18–20], several controlled trials [10, 21–23]. Because of this much variability in response, it is important to understand specifically if macroscopic cerebellar network heterogeneity is a driving factor in this variance.

Cerebellar-targeted TMS using theta-burst patterns has improved negative symptoms in limited short-term applications, such as 10 total sessions [11, 18, 19]. Further controlled trials have demonstrated that additional factors are likely at play in governing response to cerebellar TMS [10, 21–23]. These can include coil configurations, orientation, and placement, pulse waveforms, repetitive pulse patterns, the total number of pulses, and intensity [24]. Furthermore, variability in the optimization of scalp targets and protocol designs can lead to inconsistent outcomes. For example, some studies have localized the cerebellar target using scalp landmarks [12, 21], while others have employed neuronavigation tools [10, 23].

The methodological differences stated above can significantly influence results, highlighting the need for a more standardized and precise approach. As mentioned earlier, brain networks associated with the cerebral cortex are found in the cerebellum [1]. Neuroimaging has elaborated on the functional role of these connections within the cerebellum which include verbal ability, attention, memory, and emotion separate from motor regions [25]. These observations coincide with a hypothesis that schizophrenia may be better characterized as a cerebellar dysfunction of cognitive circuitry [5, 6, 26–28].

Findings from studies [5, 6, 26–28] report reduced cortical inhibition in schizophrenia [29], and network level dysregulation of the cerebello-thalamic-cortical circuit [30, 31].

Treatment outcome variability has encouraged work focused on targeting efficiency since participant-specific *functional* anatomy may be a governing principle [32, 33]. Additionally cerebellar networks are more variable across individuals as compared to cortical networks [34]. In cognitive neuroscience studies, participant-specific functional localization changed the effect size of a TMS intervention from d=0.34 to d=1.13 [33]. Individualized network-specific targeting has shown network-based responses in connectivity [35, 36] and behavior [37]. The greatest symptom improvements correlated with changes in the network connectivity between the cerebellum and DLPFC in individuals with schizophrenia further reinforcing the theory of network-level dysfunction [38]. Since there is profound variability in network distribution across individuals, there is potential for stimulation reaching an “off-target” network instead of the intended ‘on-target’ network. Thus, failure to find effects between real and sham participants [10] may be explained by stimulation variation.

The overarching hypothesis of our current study is that network activation may be required for stimulation of the cerebellum, but it is unclear which network is specifically targeted in schizophrenia. Individuals with schizophrenia may have spatial topographies of cerebellar networks or varying head morphology, which lead to differences in stimulation or impact the ability of TMS to reach the intended target. To examine the possibility that cerebellar directed TMS may reach different networks in different participants, we conducted an evaluation using neuronavigation. In our prior studies, we have indexed cerebellar stimulation by motor threshold intensities, so we modeled this stimulation intensity for coils placed on the scalp for targeting. We also collected individual resting state imaging to identify brain networks within the cerebellum. Thus, we sought to identify the source of variability in TMS network responses in these individuals.

**Figure 1.**
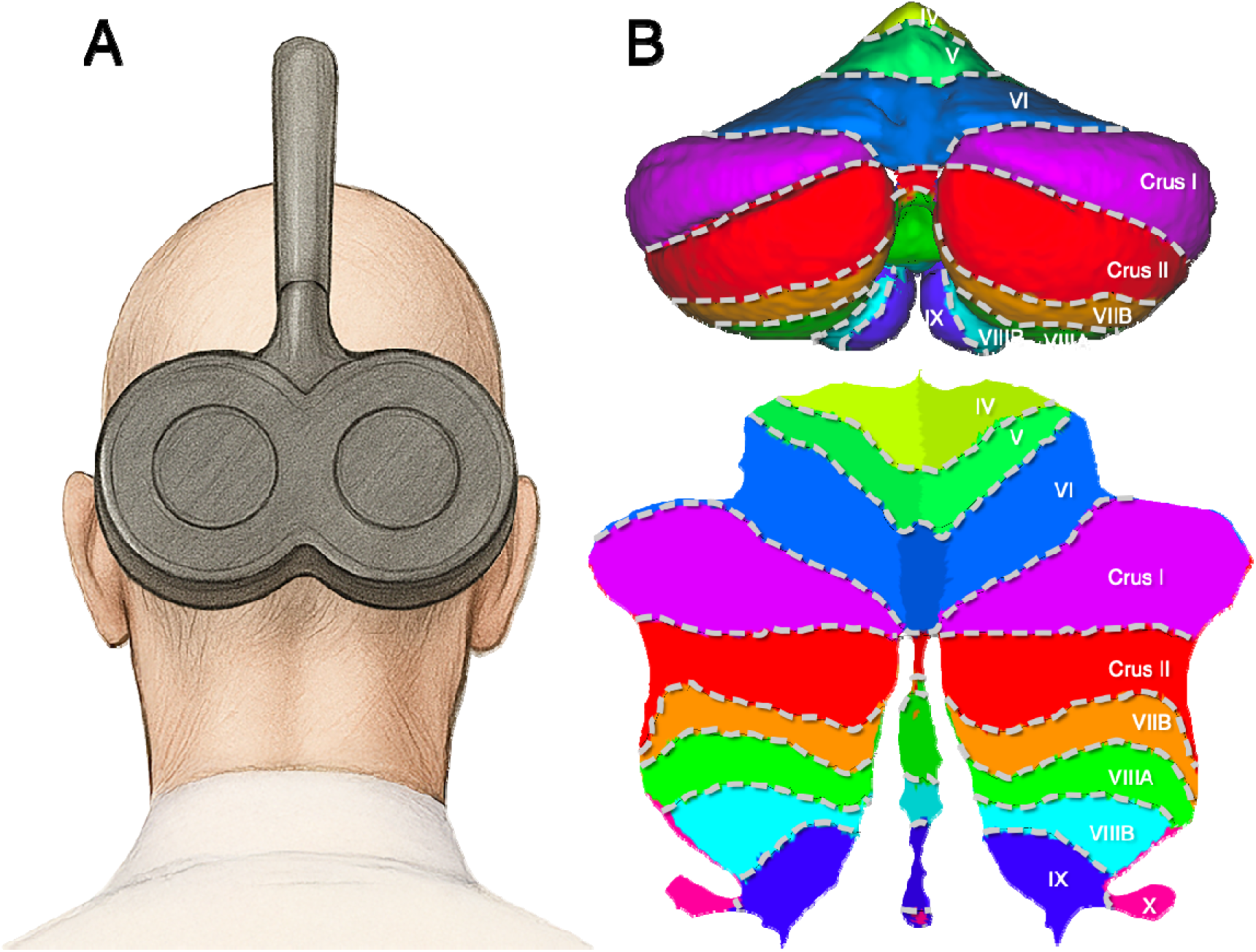
Overview of experimental methods. A. Diagram of coil position for cerebellar TMS. B. Top: Surface representation of cerebellum. Bottom: Flattened cerebellar surface with cerebellar lobules labelled.

## METHODS

### Overview

For this study, we explored the maximal network impact of cerebellar transcranial magnetic stimulation. We acquired resting state fMRI data from 27 participants with a diagnosis of schizophrenia or schizoaffective disorder.

### Participants

Participants were English-speaking adults with a schizophrenia or schizoaffective disorder. The ages ranged from 18–55 yrs (n 27, mean age 34.07 yrs, 70% male). All participants were in stable outpatient treatment with no recent (within the past 30 days) hospitalizations or changes in their medication regimens and no moderate to severe substance use disorder in the past 3 months. All participants were judged by study staff to be capable of completing the study procedures and provided written informed consent as approved by the institutional review board of McLean Hospital.

### MRI Data

All MRI data was collected at the Brain Imaging Center at McLean Hospital using the Siemens 3T Prisma scanner with a 64-channel head coil. The sequence collected structural MRI and resting state functional MRI using a modified version of the HCP Lifespan Prisma protocol [39]. All participants received a high-resolution structural MRI scan to obtain a multi-echo MPRAGE sequence [T1 MEMPRAGE (1mm^3^ voxels)] for anatomical referencing of the functional recordings. Six-minute rs-fMRI (2.3 mm^3^ voxels; 650ms TR). The initial rs-fMRI was used in combination with MRI data to isolate individual anatomical landmarks and resting-state networks for targeting rTMS modulation. During resting state sequences, participants were instructed to stay awake, to keep their eyes open and on the fixation cross being projected, and to think of whatever thoughts came to mind.

### fMRI Analysis

The Halko laboratory has developed a standardized toolbox for processing rsfMRI (and task-state fMRI during gradual-onset continuous performance task [gradCPT]). Briefly, all data is preprocessed through the field-standard toolbox of fMRIprep (Esteban et al 2019). fMRIprep performs field-inhomogeneity correction, motion-correction, and nuisance variable estimation (motion translation/rotation and derivatives, spike detection, global signal, white matter, and gray matter) using multiple software packages including routines from FreeSurfer, FSL, and CompCor (Behzadi et al., 2007). These corrected data are then registered into native participant space, MNI standard space (MNI152NLin2009cAsym), and if needed, FreeSurfer surface space (fsaverage6). With custom Python nilearn-based analysis scripts, functional connectivity is calculated by seeding a standardized 7-network brain parcellation (Yeo et al 2011). Briefly, data is temporally filtered by selecting low-frequency signals (0.009-0.08Hz), nuisance covariates of motion, global signal, white matter and CSF are removed, and mild spatial filtering (6mm FWHM) is applied to the data. Volumes with movement >0.25mm or DVARS>1.5 are ignored from the analysis. A spatial map is generated of z-transformed correlation to the mean whole-network signal, and this map is the individualized network representation of the standard networks. For analyses that require data reduction to a singular value, mean network connectivity within the network of interest is computed using the mean connectivity of all voxels within the network mask.

### E-field Modeling using SimNIBS

SimNIBS[40]long with the CHARM segmentation, was utilized to model the electric field (E-field) based on individualized MRI scans obtained for each participant. A total of N = 27 personalized head models were generated. To facilitate precise targeting for Transcranial Magnetic Stimulation (TMS), a real TMS coil was placed to target the cerebellum and the location was stored using the Brainsight neuronavigation system for each participant were used to derive participant-specific coordinates. The simulated MagVenture_Cool-B65 coil model was then placed over the recorded stimulation location for each participant to calculate the corresponding E-field distribution.

### SUIT

The SUIT toolbox [41] in MATLAB was employed to generate cerebellar networks in the native space across participants. This process involved segmenting the anatomical images into gray and white matter, followed by the isolation of the cerebellar tissue. The isolated cerebellum was then normalized using the DARTEL template. The affine matrix and deformation matrices from this normalization step were applied to resample the fMRI-derived network maps, which were subsequently resampled into cerebellar space, resulting in participant-specific cerebellar network maps in native space.

## RESULTS

### Cerebellar resting state functional network parcellations in schizophrenia spatially identical to unaffected controls

We first sought to examine the spatial relationship of cerebellar networks of the patient sample with the normative sample parcellation [1]. Participant-specific winner-take-all parcellations were generated and projected onto flat representations of the cerebellum. To create a sample-wide winner-take-all parcellation, the cerebellum vertex-wise functional connectivity values of each network were computed for each participant and then averaged, these averages were used to compute the winner-take-all vertex wise network membership (Figure 2B). From a qualitative impression, the schizophrenia sample is remarkably similar to the normative sample, given the normative sample was generated from a larger sample size (n=1000) (Figure 2A) and had regressed signals from the occipital lobe as nuisance signals (see details in [1]).

**Figure 2.**
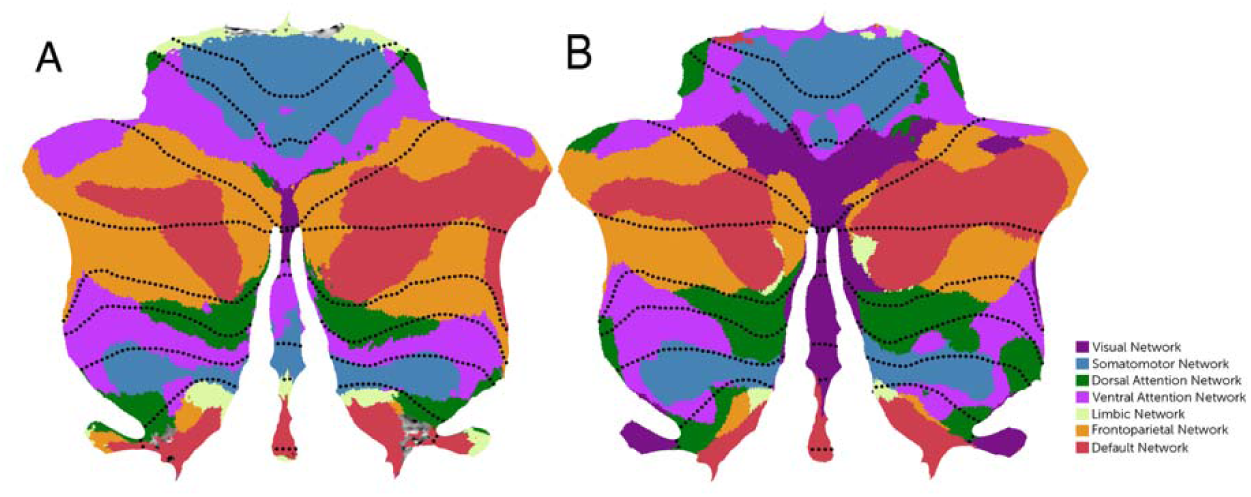
Seven network parcellations of the cerebellum. A) Reference winner-take-all cerebellar network parcellation from 1000 unaffected controls (Buckner et al 2011). B) winner-take-all cerebellar network parcellation from 27 participants with schizophrenia.

### iTBS-Induced Electric Field Peaks Localized to Vermis and Bilateral Crus II

TMS field efficacy on the cerebellum is completely unknown, particularly for vermal stimulation in schizophrenia. To assess the topographic distribution of the TMS field, we determined the resting motor threshold for each participant and then placed a TMS coil targeting the cerebellar vermis and recorded the coil position in individual anatomical space. Simulations of stimulation at 100% of the resting motor threshold were computed with SimNIBS. Each field map was then normalized into SUIT atlas space, and the mean field was computed across all participants (Figure 3). Qualitatively, the EF peaks denoted by values above 19 V/m were found to be predominantly distributed around vermis and bilateral crus regions. Although there is no clear agreement on the exact strength of the electric field (EF) required to produce an electrophysiological effect, empirical evidence indicates that an EF ranging from 20 V/m to 50 V/m is likely to be effective. [42]

**Figure 3.**
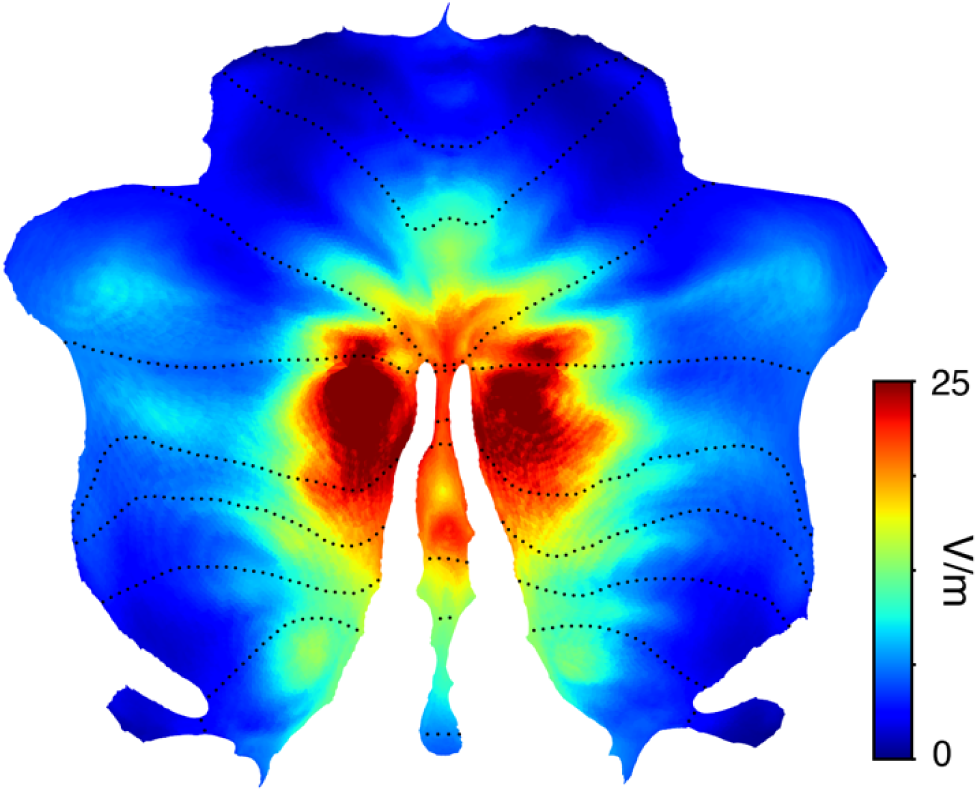
Mean TMS induced E field distributions in 27 participants with schizophrenia.

### Functional networks exhibited variability at the individual participant level

All individuals did not have the peak field intensity in the same network. This variability across participants could have many sources, therefore we examined individual network topographies to appreciate these findings. Figure 4 highlights some of these results, which suggests that the variability in network receptivity of TMS is likely due to individual differences in network topology and not due to individual differences in field distributions on the cerebellum. We note that these differences are likely due to the selection of a ‘winner-take-all’ parcellation approach which highlights these effects.

**Figure 4.**
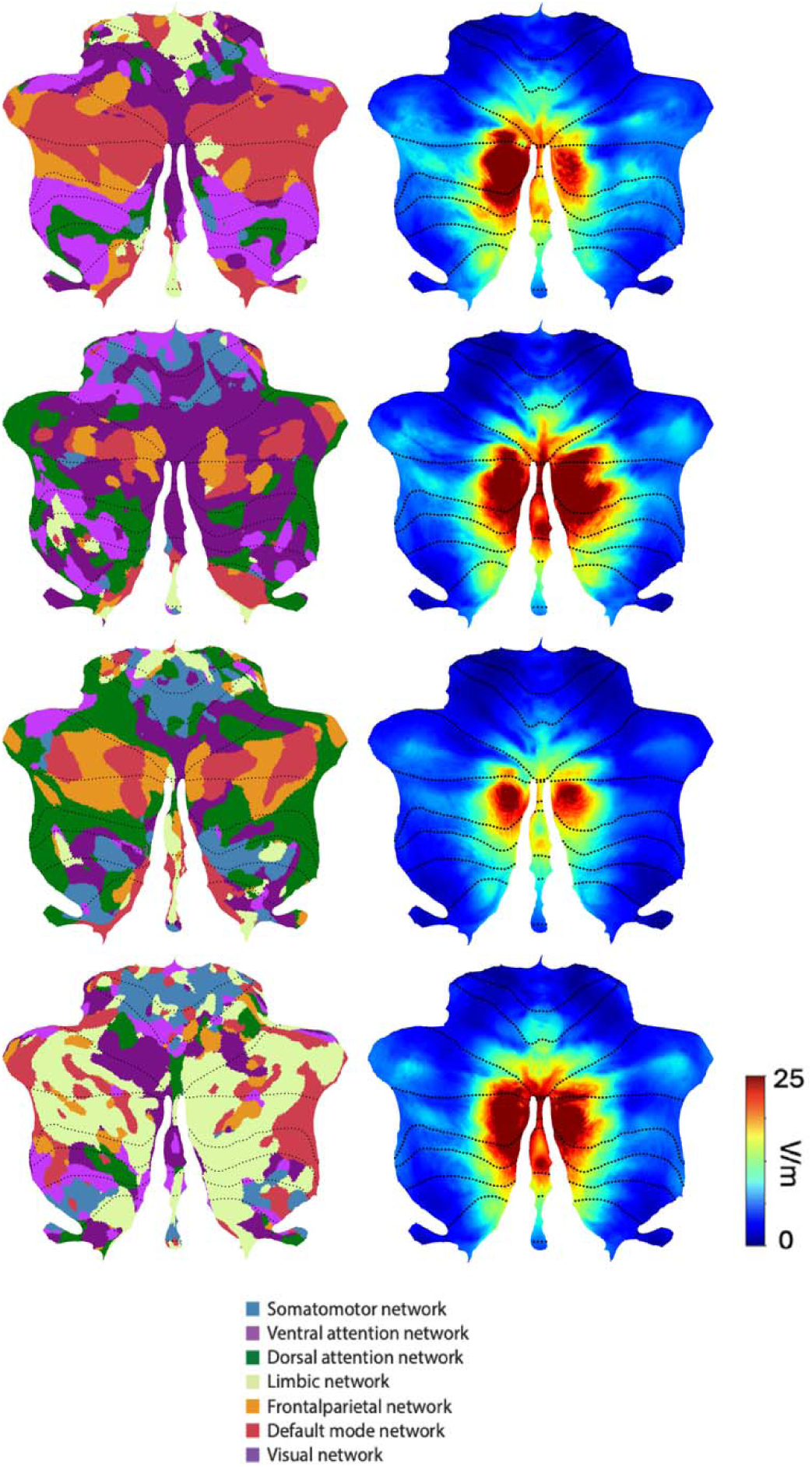
Representative examples of individual participant parcellations and electric field maps. Left column: Individualized winner-take-all seven network parcellation of the cerebellum based on individual participant-specific network functional connectivity. Right column: Simulated electric field on the cerebellum for individualized coil placement and TMS stimulation intensity. A. Participant for whom peak field is in default network. B. Participant for whom peak field is in frontoparietal network. C. Participant for whom peak field is in frontoparietal network. D. Participant for whom peak field is in limbic network.

### Predominant Engagement of Cognitive Networks by Vermis-Targeted iTBS Across Participants

Having established the TMS field reaches the cerebellum, we next sought to determine which networks of the cerebellum are impacted by stimulation. It is well understood that normative network organization may not reflect individual organization, so we first generated participant-specific cerebellar network parcellations and compared that organization to individual electrical field distributions in the cerebellum. In electrical simulations, local inhomogeneities in meshes can result in abnormally high peak values. To ensure that we found the locations of maximum field, we identified the locations of the 99^th^ percentile of the electric fields across the entire cerebellum. For each participant, we identified the location of peak stimulation (99th percentile) and determined which cerebellar network that location belonged to. This was to identify which network receives the strongest TMS effect. Figure 5 summarizes these results. Across participants, ANOVA reveals differential network impact of vermal cerebellar TMS (F(27,6) = 28.56, p<0.01), demonstrating that TMS predominantly will impact the limbic network, the frontoparietal control network and the default network (Figure 5B). The peak intensity was found in the default network in the majority of participants, followed by the frontoparietal control network and limbic network (Figure 5A).

**Figure 5.**
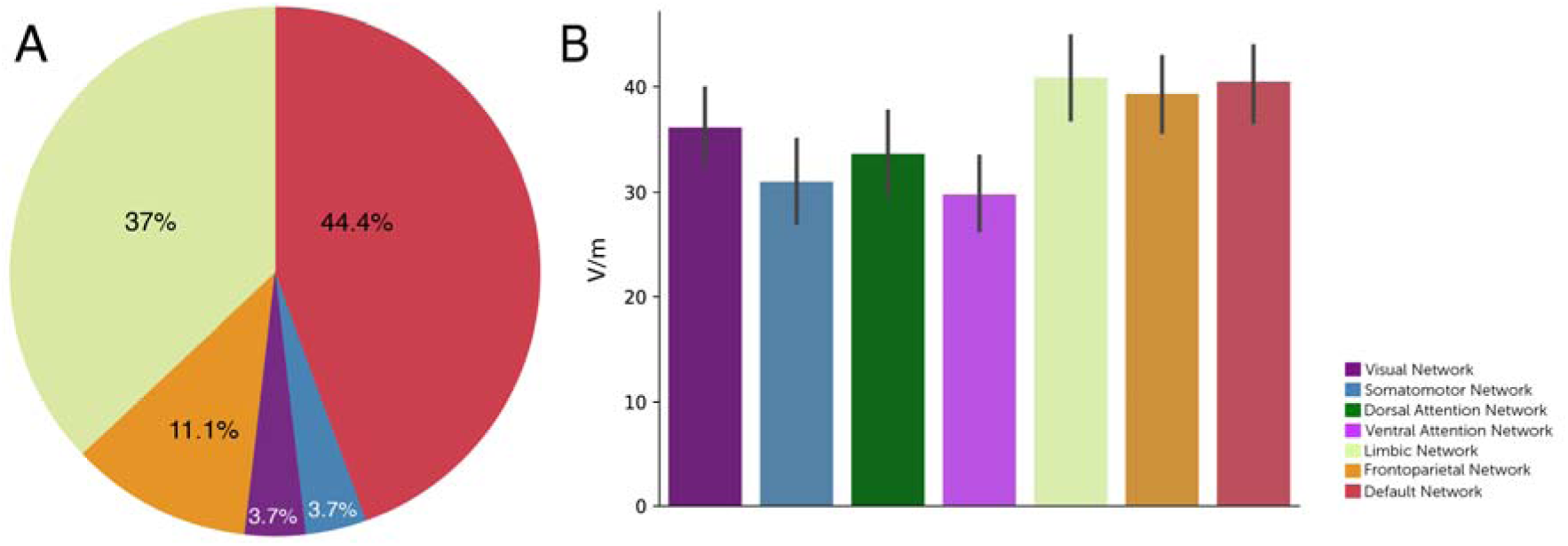
Location and network distribution of simulated TMS electric field. A. Network membership of peak fields locations in cerebellum. Each participant was assigned a network based on the location of their peak electrical field within their own individualized cerebellar network parcellation. B. Average of peak field by network across participants.

## DISCUSSION

The increasing number of clinical trials investigating cerebellar TMS as an adjuvant treatment strategy for schizophrenia motivated our investigation of the targeted brain-network effects of cerebellar TMS. We collected data from real TMS participants to produce an accurate simulation of the TMS field from a standard figure of 8 coil on the cerebellum. We combined this information with individual functional connectivity analyses of cortical-cerebellar networks. Our study found that TMS reaches the cerebellum, but the intensity of stimulation varies across participants. Moreover, we found that the regions of the cerebellum attained by TMS primarily correspond to associative and affective networks of the default network, fronto-parietal control, and limbic networks. This result is largely consistent with cerebellar stimulation effects observed in clinical and healthy populations [36–38].

Our results may explain why there is such variability in response in cerebellar targeted TMS in schizophrenia. We observed that many of the networks associated with cognition and emotion are represented in the cerebellar superficial midline structures that TMS can reach, and within participants, these representations may be variable. This suggests that that varying anatomy and physiology may influence clinical effects. We observed several sources of this variability. First, motor threshold varies across participants, when cerebellar rTMS is indexed based on cortical motor excitability, it is reasonable to conclude that variability in motor threshold indexed stimulation intensity on the cerebellum will create variable fields upon the cerebellum. Variability in the tissue of the neck and head may also cause changes in the electrical field in the cerebellum where some individuals may have an increased distance between cerebellum and feasible coil placement. Finally, cerebellar neuroanatomy also plays a role in individual electrical field distributions, as the formation of the cerebellum may present easier or more difficult access to non-invasive brain stimulation. All these sources of variance are summed together to create the variability of electrical field distribution upon the cerebellum.

To successfully target the cerebellum and adjust electrical field intensity to match the effective field, it is important to also consider individual network organization. The efficacy of TMS depends on functional localization of a target – holding a coil near a target may still reach the intended target when increased intensity is applied. Thus consideration of intensity must be considered alongside the consideration of the target location. To date, it is unknown which network target should be functionally determined for TMS efficacy. In a previous study, we demonstrated the identification of a cerebellar prefrontal “circuit” that is associated with negative symptom severity and showed that restoring functional connectivity between those two nodes with TMS improved symptoms[23]. However, it is not established if other networks are involved in other symptoms, or if this same circuit governs all symptoms. Recent work from our group and others has highlighted functional connectivity deficits within the cerebellum that relate to symptoms [10, 43–45]. Our goal in the present study was not to identify which network is dysfunctional, but rather to establish what networks are targeted with non-invasive cerebellar stimulation. Our analysis revealed that the impact of cerebellar TMS is likely spread across several networks (default network, frontoparietal control, and limbic) and that individuals may have varied connectivity profiles that highlight one network over another, thus resulting in multiple different network effects. This may be an additional source of variance for clinical effects from cerebellar stimulation.

Our findings support a hypothesis that vermal-targeted TMS likely reaches the cerebellar vermis and midline hemispheric structures at the level of Crus I/II. By utilizing real TMS values in our modeling and participant-specific accurate head models, we found substantial variability in individual participant electrical field. We hypothesize that incorporating magnetic field modeling may be critical to understanding TMS response variability. Similarly, this approach may be useful prospectively to titrate the appropriate TMS intensity to create a homogenous cerebellar field across participants. To date, such an attempt has not been made. Some studies have examined the variability of TMS applied to the cerebellum as a modeling question [46], and others have adopted strategies to adjust the intensity by tissue distance [47]. However, these approaches fail to consider the functional organization of the cerebellum. Adjusting intensity for the nearest tissue will not necessarily improve stimulation efficacy, as one must also consider the specific networks present in the target functional region.

This study is among the first to explore the effects of intermittent theta burst stimulation (iTBS) on the cerebellum using electric field modeling in utilizing cerebellar-specific neuroimaging tools. We utilized real-world data from a relevant target population, having real coil positioning, real motor threshold determinations, and observed di/dt stimulation intensity at the coil surface. We also demonstrate the utility of examining individual functional connectivity parcellations as a source of variability of TMS effects.

Several limitations of the study should be acknowledged. One participant presented with a misaligned cerebellar position due to the presence of a localized cyst. Although coil placement and electric field modeling were adjusted to accommodate this anatomical variation, the case highlights the importance of evaluating iTBS precision on a case-by-case basis- further emphasizing the need for personalized stimulation protocols. As we took a personalized real-world approach, larger sample sizes could not be collected. The approaches contained here may be virtually extended into larger datasets where TMS information is unknown but could be estimated, and such work is urgently needed. Future work could utilize these findings to provide prospective information for TMS practitioners to determine optimal TMS settings and protocols for efficacious treatment.

Despite these factors, the central aim of this study was to establish the presence and spatial distribution of local electric fields (EF) following iTBS. We aimed to assess and report the feasibility of this approach and for conducting this investigation to a larger population. In doing so, we intentionally kept several factors uncontrolled to better reflect the real-world conditions under which neuromodulation therapies are practically delivered. Also, we argue that the confounding variables are unlikely to substantially impact the core findings related to EF localization and its alignment with functional cerebellar networks.

There is an increasing body of evidence underscoring the importance of personalized treatment approaches. This study contributes to this growing understanding by providing insights into inter-individual variability in cerebellar network activity. Specifically, it illustrates the spatial pattern of local electric fields produced by intermittent Theta Burst Stimulation (iTBS) at the cerebellar vermis and highlights the cerebellar networks that are most effectively engaged by this intervention. The findings from this modeling study reinforce the critical need to evaluate both the structural and functional pathophysiological characteristics of the cerebellum before selecting target locations for auditory hallucinations in schizophrenia, maximizing therapeutic benefit and symptom relief.

## Data Availability

All data for the present study are deposited in the NIMH Data Archive (NDA: https://nda.nih.gov).

## ACKNOWLEDGEMENTS

This study was supported by NIH awards R01MH126000, R01MH111868, R56MH125995 and R01MH116170.

